# Assessment of Potential Risk Factors for COVID-19 among Health Care Workers in a Health Care Setting in Delhi, India - A Cohort Study

**DOI:** 10.1101/2022.02.28.22271674

**Authors:** Mridu Dudeja, Aqsa Shaikh, Farzana Islam, Yasir Alvi, Mohammad Ahmad, Varun Kashyap, Vishal Singh, Anisur Rahman, Meely Panda, Neetushree, Shyamasree Nandy, Vineet Jain

## Abstract

**Introduction:** Health care workers (HCW) are among the most vulnerable for contracting the COVID-19 infection. Understanding the extent of human-to-human transmission of the COVID-19 infection among HCW is critical in management of this infection and for policy making. We did this study to observe seropositivity and estimate new infection by seroconversion among HCW and predict the risk factors for infection.

**Methods:** A cohort study was conducted at a tertiary dedicated COVID-19 hospital in New Delhi during first and second wave of the COVID-19 pandemic. All HCW working in the hospital during the study period who come in contact with the patients, were our study population. The data was collected by a detailed face to face interview along with serological assessment for anti-COVID-19 antibodies at baseline and endline, and assessment of daily symptoms. Prediction of potential risk factors for seroprevalence and seroconversion was done by logistic regression keeping the significance at p<0.05.

**Results:** A total of 192 HCW were recruited in this study, out of which, 119 (61.97%) at baseline and 108 (77.7%) at endline were seropositive for COVID-19. About two-third (63.5%) had close contact, 5.2% had exposure during aerosol procedures, 30.2% had exposure with a patient’s body fluid while majority (85.4%) had exposure to contact surface around the patient. Almost all were wearing PPE and following IPC measures during their recent contact with a COVID-19 patient. Seroconversion was observed among 36.7% of HCWs while 64.0% had a serial rise in titer of antibodies during the follow-up period. Association of seropositivity was observed negatively with doctors [OR:0.353, CI:0.176-0.710], COVID-19 symptoms [OR:0.210, CI:0.054-0.820], comorbidities [OR:0.139, CI: 0.029 - 0.674], and recent Infection Prevention Control (IPC) training [OR:0.250, CI:0.072 - 0.864], while positively associated with partially [OR:3.303, CI: 1.256-8.685], as well as fully vaccination for COVID-19 [OR:2.428, CI:1.118-5.271]. Seroconversion was positively associated with doctor as profession [OR: 13.04, CI: 3.39 - 50.25] and with partially [OR: 4.35, CI: 1.070 - 17.647], as well as fully vaccinated for COVID-19 [OR: 6.08, CI: 1.729 - 21.40]. No significant association was observed between adherence to any of the IPC measures and PPE (personal protective equipment) adopted by the HCW during the recent contact with COVID-19 patients and seroconversion.

**Conclusion:** A high seropositivity and seroconversion could be either due to exposure to COVID-19 patients or concurrent immunization against COVID-19 disease. In this study the strongest association of seropositivity and seroconversion was observed with recent vaccination. IPC measures were practiced by almost all the HCW in these settings, and thus were not found to be affecting seroconversion. Further study using anti N antibodies serology, which are positive following vaccination may help us to find out the reason for the seropositivity and seroconversion in HCW.

## INTRODUCTION

COVID-19 belongs to the large family of viruses *Coronaviridae*, that cause a wide spectrum of symptoms and diseases in human beings. However, these viruses have the peculiar property to constantly change and become diverse, which has helped them spread, survive and puts our health system into jeopardy. COVID-19 transmits from one host to another host via respiratory droplets, aerosol, contact with bodily fluids and with contaminated surfaces (1). Individuals who are asymptomatic may be able to transmit infection, while individuals who have not reported close proximity to any known case have also been infected(2)(3)

As a country with multifarious ethnic diversity and cultural assortment, India’s stand on COVID-19 has been proactive. India has the largest number of confirmed cases in Asia, and the second highest number of confirmed cases in the world(4). Available from: https://covid19.who.int} With 3.06 crore cases, the recovery rate is 2.98 crore which implies that the case fatality rate (CFR) is relatively low at 1.49% as against the global values of 4.7%(5). So far thousands of healthcare workers; including doctors, nurses and paramedical staff have succumbed to COVID-19 in India. The national capital being the nucleus of the strategic amalgamation of all policies and stratagem stirs a discrete figure. Test and treat policy is the main backbone of Delhi’s fight against the COVID-19. Second wave saw an increase in cases and deaths mostly in a phased wise manner; waning and waxing off after a sudden short and high peak.

Health workers are at the heart of the crisis which the world is experiencing. They are facing acute challenges while treating patients and at the cost of their own health and that of their families. Besides the increased psychological burden, due to the heightened patient care burden and lack of empathy; their overall wellness is something which is the need of the hour.(6). Starting from, line listing, diagnosis, treatment, rehabilitation, home visits to prevention; all of those require the health work force to plunge into as front liners. To add to these sufferings are the financial insecurities, violence, wrath of families affected and governmental mismanagements. HCW providing COVID-19 care are at increased risk of acquiring infection if there is slightest breach in personal protection. They are valuable and scarce resources who cannot be spared for getting isolated for treatment and quarantine. Their health in terms of disease status and mentation besides being a concern for themselves also affects the hospital service delivery. The WHO reported that one in ten health workers is infected with the virus. Infection is more common among nursing staff while death is seen more among doctors, with highest case fatality rate seen in age group over 70 years (7). They also have a role in the implementation of adequate IPC measures in health care facilities. Several advisories and directives have been updated by the Ministry of Health and Family Welfare EMR Division, for managing this crisis among Health care workers. They stressed for activating Hospital Infection Control Committee (HICC) and identifying nodal officer to respond to Health Associated Infections (HAIs) and following updated guidelines.

Investigating the extent of for COVID-19 infection and assessing the potential risk factors among health workers is essential for characterizing virus transmission patterns, preventing future infections of health workers and preventing the health-care-associated spread of COVID-19. With the emergence of mutant forms and the rising disarray between the health system and the political clutter, it’s a priority to safeguard the frontliners and make them battle ready.

So, we did this study with the objective of (1) to understand the extent of human-to-human transmission of COVID-19 by estimating the seroconversion among HCW in recent contact with a COVID-19 patient; (2) to characterize the range of clinical presentations of infection (3) to evaluate the effectiveness of infection prevention and control (IPC) measures among HCW; and (4) to find out risk factors for seroconversion and serological response.

## MATERIAL AND METHODS

### Study design and population

This was a prospective cohort study carried out among health care workers (HCW) between December 2020 to June 2021, the period covered India’s intense second wave of COVID-19 pandemic.

The study was conducted at Hamdard Institute of Medical Sciences and Research (HIMSR) and Hakeem Abdul Hameed Centenary Hospital (HAHC), which is a dedicated COVID-19 Hospital of 200 beds, located in South East Delhi, India. The study population included all the health personnel who were working in this hospital and had come in contact or been exposed recently to a COVID-19 patient receiving care.

#### Inclusion criteria

All HCW exposed to COVID-19 patient receiving care in this health care facility within 72 hours of confirmation of the diagnosis with exposure either to

- Close contact (within 1 m) to laboratory-confirmed case
- or exposed to case’s blood or body fluids,
- or exposed to case’s used materials, devices or equipment,
- or environmental surface around case including his/her bed, table, wheelchair, ward corridor etc.

They were enrolled irrespective of their use of Personal Protective Equipment (PPE), any symptoms, and vaccination status.

#### Exclusion criteria

- HCW who also works in another healthcare facility
- HCW who had already suffered from COVID-19 before the start of the study
- HCW who are COVID-19 infected or have a confirmed COVID-19 case among their household/close contacts.
- HCW who are so clinically serious that they cannot participate in the study.

### Sample Size

For determining sample size, we used methods of Kelsey, Fleiss and Fleiss with continuity correction. The ratio of unexposed to expose is kept as 1:1. We hypothesize a 36% outcome in the unexposed group based on the previous studies (8–10). With a risk ratio of 1.7 and Odds ratio of 2.8, we got the sample size of 138, which was increased to 180 considerating the attrition rate at 25%.

### Definition

- Healthcare worker: Any staff in the healthcare facility involved in the provision of care for a COVID-19 infected patient. It included those who have been present in the same area as the patient as well as those who may not have provided direct care to the patient but who have had contact with the patient’s body fluids, potentially contaminated items or environmental surfaces.
  - Category I – All doctors like teaching faculty, residents, demonstrators and medical interns.
  - Category II – Nurses and nursing assistants
  - Category III – Lab assistants, technicians, field workers, housekeeping, sanitation workers, security personnel, general duty attendants, pharmacist, reception,
- Work place: Work place was divided into two
  - High risk Area: Areas where the risk of COVID-19 infection was high such as the emergency, aerosol generating procedure rooms, COVID-19 wards, ICU, labor room and testing center.
  - Low Risk area: Areas where there is a low risk of COVID-19 infection such as the General OPD, offices, laboratories, reception, enquiry counter and security posts.
- Type of exposure
  - Close contact exposure (within 1 meter)
    - Prolonged face-to-face exposure (> 15 minutes)
    - Exposure during aerosolizing procedures
    - Direct Exposure with body fluid
  - Patient’s materials exposure: (personal belongings, linen and medical equipment)
    - Exposure to Patient’s body fluid via materials exposure
  - Surface exposure:
    - Exposure to Patient’s body fluid via surface around patient

### Study Outcome

#### SEROPOSITIVITY

Study participants who were positive (A/C.O. &#x2265;1) for SARS-CoV-2 antibodies detected with WANTAI SARS-CoV-2 Ab ELISA at Baseline. This included all those who were positive due to prior COVID-19 infection or as a result of vaccination against the COVID-19.

#### SEROCONVERSION

Study participants who were negative (A / C.O. < 1) for SARS-CoV-2 antibodies detected with WANTAI SARS-CoV-2 Ab ELISA at baseline but became positive (A / C.O. >1) in the end line serum sample collected between 22-28 day.

RISING TITRE: Rise in titre for SARS-CoV-2 antibodies detected with WANTAI SARS-CoV-2 Ab ELISA at endline from baseline

#### SECONDARY INFECTION RATE

Those HCW who were enrolled after the resent exposure with COIVD 19 patients and confirmed for new infections of COVID-19 assessed through serological assays on paired samples divided by susceptible contacts (total enrolled HCW). Due to limitation of our study, it would be same as seroconversion rate.

#### SERIAL RISE IN TITRE

We considered this when the HCW was positive in the end line serum sample collected after 22-28 day and had either increased in titre (A / C.O) or had maximum possible value at both at baseline and at 22-28 day.

### Participant Recruitment

Once a confirmed case of COVID-19 was detected, identification of healthcare workers who came in contact with the patient or their contaminated objects was done by the Covid Surveillance Unit by interviewing patient and/or their attendant(s) were to understand their movement history in the hospital and to do tracing of the contacts in the hospital in the past 72 hours. The identified HCW contacts were called and informed about their potential contact and about the ongoing study. They were provided with Patient Information Sheet. If they agreed to be a part of the study, then written informed consent was obtained. Participants were also recruited through self-reporting of contacts and referrals. All details were maintained in master register of the project. (Figure 1)

**Figure 1.**
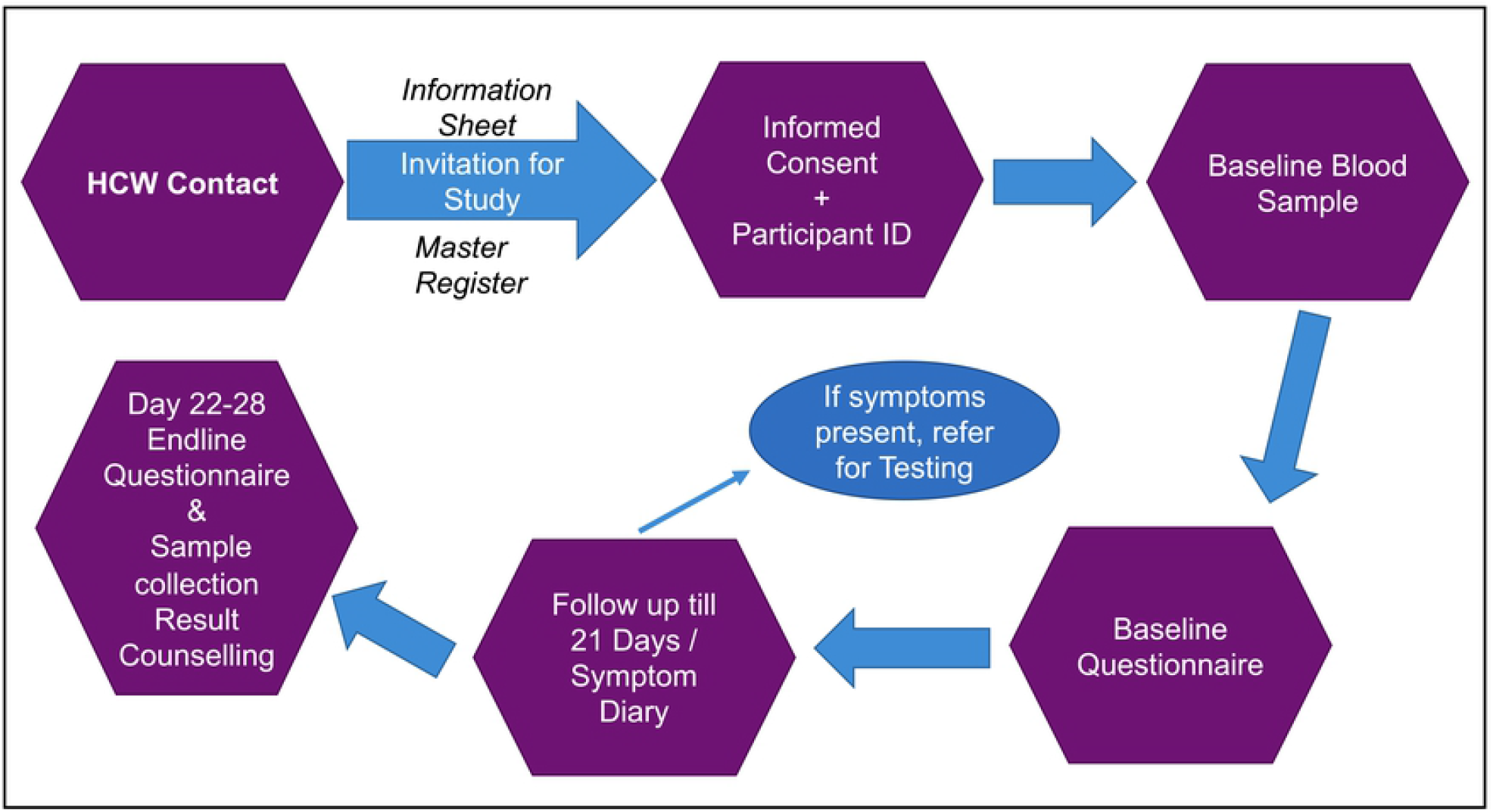
Participant flow diagram.

All HCW recruited into the study completed a researcher-administered, translated, questionnaire at baseline which covered:

- socio-demographic information
- training on infection prevention and control measures
- adherence to infection prevention and control measures and
- contact with, and possible exposure to the COVID-19-infected patient following their admission to the health care facility
- status of vaccination

The completed questionnaire was entered in Microsoft excel sheet and checked by a supervisor against the hard copy for accuracy of data entry. In additions, a baseline serum sample was collected from all study participants to check for the presence of COVID-19 antibodies.

The participants were provided with a symptom’s diary having common symptoms of COVID-19 for self-administration during the 21 days follow-up period. The participants were regularly followed up to check the filling of symptoms diary. The endline visit was scheduled at 22-28 days from the first visit during which endline assessment and second serum sample was collected. The serum samples were tested for antibodies against COVID-19 using the WANTAI serological testing kit. A value of above 1 was considered as positive. These paired serological samples allowed for detection of seroconversion, for better understanding of the secondary infection attack rate.

### Sample collection

Two millilitre of blood was collected by venepuncture from all healthcare workers who were enrolled in the study. The first sample collected after enrolment was considered as the ‘baseline blood sample’. All the subjects were recalled after 21 days (from the date of baseline sample collection) for the collection of endline blood sample. (Protocal deviation 1: In initial proposal, we planned to call only those who were sero-negative at baseline, later after protocol deviation all HCW were called for endline). The paired samples testing protocol helped in detection of asymptomatic carriers and understand the pattern of seroconversion.

The sample collected in appropriate and labelled blood vial was allowed to stand upright for 30 minutes at room temperature followed by centrifugation at 2500 rpm for 5 minutes. The samples are then sent to testing laboratory by placing the vials in a carrier box in upright position. The blood collection staffs were well trained in safe specimen handling practices and spill decontamination protocols and used appropriate PPE during the sample collection process.

### Anti-SARS-CoV-2-total antibody detection

Wantai SARS-CoV-2-Ab ELISA kit detects total antibodies against SARS-CoV-2 virus and is based on the principle of two-step incubation antigen “sandwich” enzyme immunoassay. Briefly, 100 ml of patient’s serum is added to polystyrene microwell strips pre-coated with recombinant SARS-CoV-2 antigen. Three wells are marked as negative calibrator and 2 wells as positive calibrator. 50 ml of negative and positive calibrator are added to respective wells and the plate is incubated at 37^0^C for 30 minutes. Post incubation the wells were washed 5 times with diluted wash buffer. 100μl of HRP-Conjugate was then added to each well and the pate was incubated at 37^0^C for 30 minutes. The wells were washed again washed 5 times and 50μl of Chromogen Solution A and then 50μl of Chromogen Solution B was added into each well. The plate was then incubated at 37°C for 15 minutes in dark. 50μl of Stop Solution was added into each well and mixed gently. Absorbance was measured using PR4100 microplate reader, Bio-Rad, USA (dual filter) with reference wavelength at 600∼650nm. Cut-off value (C.O.) was calculated as C. O= Nc + 0.16 (Nc = the mean absorbance value for three negative calibrators). The tested serum samples were stored at −80^0^C with proper labelling(11).

### Test result Reporting

Negative results were reported for specimens with absorbance (A) less than the Cut-off value, which meant that no SARS-CoV-2 antibodies were detected with WANTAI SARS-CoV-2 Ab ELISA (A / C.O. < 1). Specimens with absorbance equal to or greater than the Cut-off value were considered positive, which indicated that SARS-CoV-2 antibodies was detected using WANTAI SARS-CoV-2 Ab ELISA A / C.O. &#x2265; 1). All the study participants were provided with their baseline and endline antibody results and counselling was done depending on the results.

## STATISTICAL ANALYSES

Data was collected and managed in Microsoft excel with appropriate coding and later cleaned for any possible errors. The questionnaires were checked if they were complete and consistent. For analysis SPSS (version 26) was used. The frequency tests were performed after determining clear values for the outcomes. Categorical data were presented as percentage (%). Pearson’s chi-square and bivariate logistic regression was done to evaluate the independent associations of multiple factors. All tests are performed at a 5% level of significance, and thus the p value less than 0.05 (p value < 0.05) was taken as significant association.

## ETHICAL CONSIDERATIONS

Ethical considerations for doing the study were undertaken and all norms of confidentiality, autonomy, beneficence and consent were taken care of. Study started after approval by the Research Project Advisory Committee (RPAC) and Institutional Ethics committee (IEC) of HIMSR. Other necessary permissions from hospital and medical college were obtained. Written informed consent in English / Hindi was obtained from each participant before their enrolment in the study. All the project staff were trained in and followed Good Clinical Practice. All the participants who needed RTPCR test were offered the same by Hospital. All the participants with poor IPC practices were recommended for refresher IPC training

## RESULTS

Out of a total of 405 HCW approached, 192 were recruited in this study. All of them were interviewed and blood sampling for serology was done at the baseline visit. Out of them, 139 were also included at end line serology assessment, reason of lost to follow up are highlighted in Figure 2.

**Figure 2.**
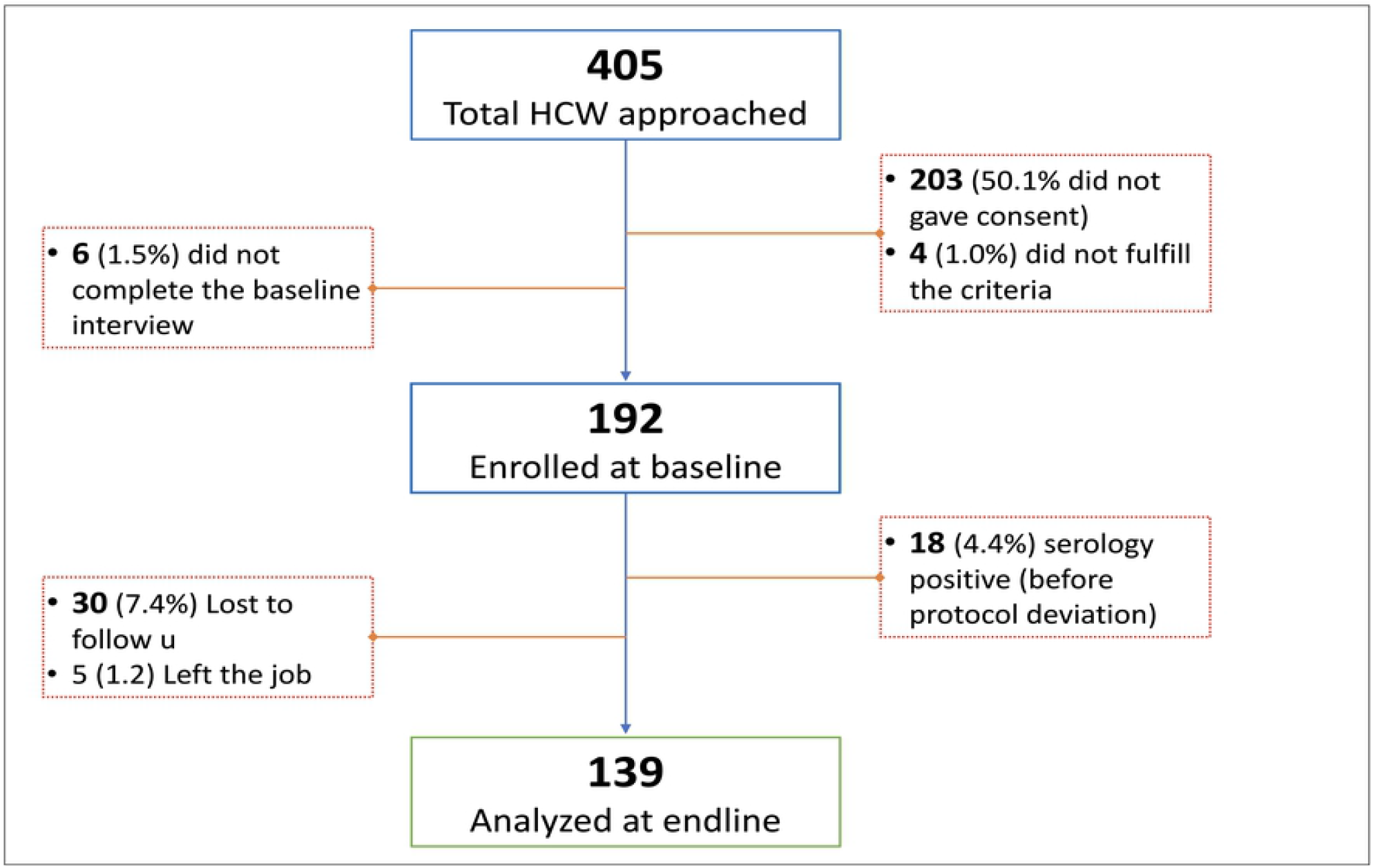
Flowchart of participants enrolment.

Sociodemographic profile, working condition, comorbidities and other characteristics of the study participants are highlighted in the table 1. Majority of the participants belongs to younger age group with mean age of 31.7 ±9.3 years with an almost equal distribution of both sexes.

**Table 1:**
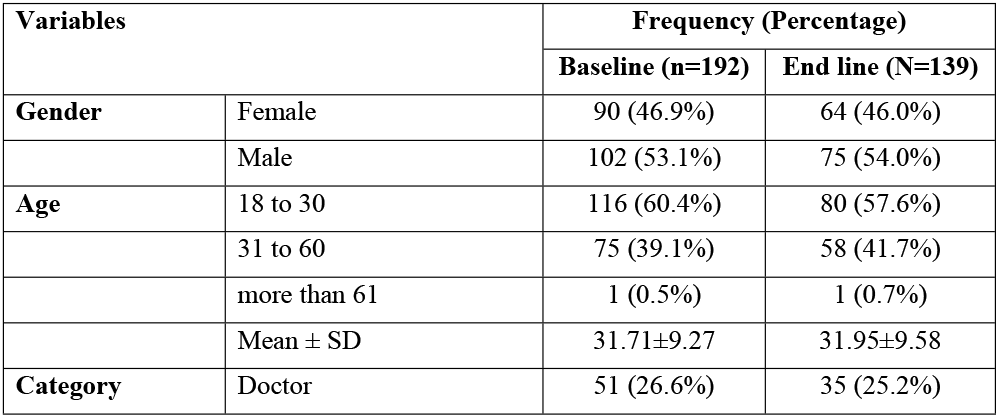
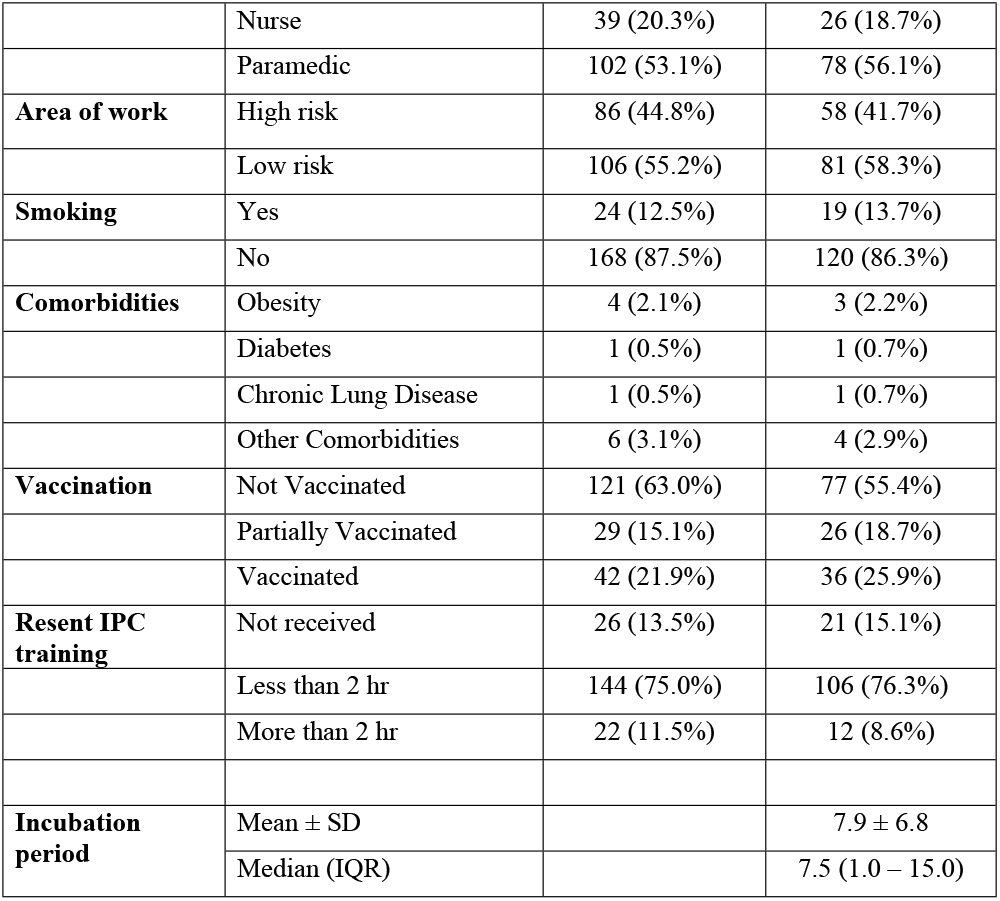
Healthcare Workers Characteristics at Baseline and Endline.

| Variables |  | Frequency (Percentage) |  |
| --- | --- | --- | --- |
|  |  | Baseline (n=192) | End line (N=139) |
| Gender | Female | 90 (46.9%) | 64 (46.0%) |
|  | Male | 102 (53.1%) | 75 (54.0%) |
| Age | 18 to 30 | 116 (60.4%) | 80 (57.6%) |
|  | 31 to 60 | 75 (39.1%) | 58 (41.7%) |
|  | more than 61 | 1 (0.5%) | 1 (0.7%) |
| | Mean ± SD | 31.71±9.27 | 31.95±9.58 |
| Category | Doctor | 51 (26.6%) | 35 (25.2%) |
|  | Nurse | 39 (20.3%) | 26 (18.7%) |
|  | Paramedic | 102 (53.1%) | 78 (56.1%) |
| **Area of work** | High risk | 86 (44.8%) | 58 (41.7%) |
|  | Low risk | 106 (55.2%) | 81 (58.3%) |
| **Smoking** | Yes | 24 (12.5%) | 19 (13.7%) |
|  | No | 168 (87.5%) | 120 (86.3%) |
| **Comorbidities** | Obesity | 4 (2.1%) | 3 (2.2%) |
|  | Diabetes | 1 (0.5%) | 1 (0.7%) |
|  | Chronic Lung Disease | 1 (0.5%) | 1 (0.7%) |
|  | Other Comorbidities | 6 (3.1%) | 4 (2.9%) |
| **Vaccination** | Not Vaccinated | 121 (63.0%) | 77 (55.4%) |
|  | Partially Vaccinated | 29 (15.1%) | 26 (18.7%) |
|  | Vaccinated | 42 (21.9%) | 36 (25.9%) |
| **Resent IPC training** | Not received | 26 (13.5%) | 21 (15.1%) |
|  | Less than 2 hr | 144 (75.0%) | 106 (76.3%) |
|  | More than 2 hr | 22 (11.5%) | 12 (8.6%) |
| **Incubation period** | Mean ± SD | | 7.9 ± 6.8 |
|  | Median (IQR) |  | 7.5 (1.0 – 15.0) |

More than half of them were paramedic and a quarter were doctor and nurse. About 63% were unvaccinated at baseline which was least among doctors. (Figure 3). All the HCW had taken Covishield (AstraZeneca/ChAdOx1 nCoV-19) vaccine. We found incubation period of 7.9 ± 6.8 days in our study population. Median incubation period was also similar 7.5 days as shown in Table 1

**Figure 3:**
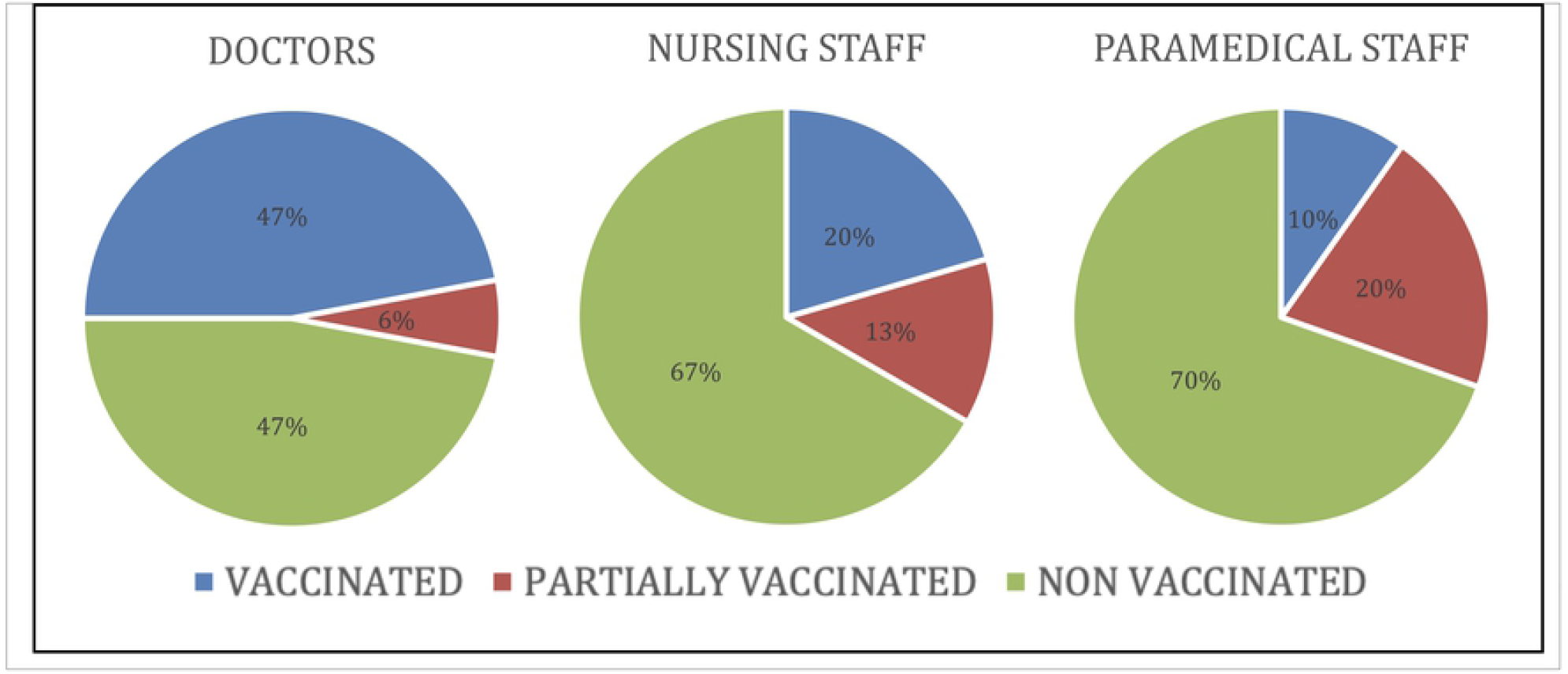
Vaccination status among HCW at baseline.

About two third (63.5%) of the HCW had close contact exposure with COVID-19 patient, and among them 27 had face to face prolong exposure and 10 had exposure during aerosol generating procedure while exposure with body fluid was observed in 10.7%. Exposure with patient material was seen in 30.2% while exposure with surface around patient was observed in 85.4%. Exposure to patient’s body fluid via patients’ material and via surface around patient was also seen in 2.1% and 2.6% respectively. (Table 2)

**Supplementary Table 1:** Healthcare Workers’ Exposure across different profession.

| Exposure | Healthcare Worker Category | Frequency | Percent |
| --- | --- | --- | --- |
| Close contact exposure<br>n= 122 | Doctors (51) | 35 | 68.6 |
|  | Nursing Staff (39) | 31 | 79.5 |
|  | Paramedical Staff (102) | 56 | 54.9 |
| Prolonged Face to face<br>exposure n=27 | Doctors | 8 | 15.7 |
|  | Nursing Staff | 7 | 17.9 |
|  | Paramedical Staff | 12 | 11.8 |
| Exposed to aerosol<br>generating procedure n=10 | Doctors | 4 | 7.8 |
|  | Nursing Staff | 4 | 10.3 |
|  | Paramedical Staff | 2 | 2.0 |
| Direct contact with patient's<br>body fluids n=13 | Doctors | 5 | 9.8 |
|  | Nursing Staff | 5 | 12.8 |
|  | Paramedical Staff | 3 | 2.9 |
| Exposure with Patient's<br>materials n=58 | Doctors | 15 | 29.4 |
|  | Nursing Staff | 12 | 30.8 |
|  | Paramedical Staff | 31 | 30.4 |
| Exposure with patient's body<br>fluids via Patient's materials<br>n=4 | Doctors | 3 | 5.6 |
|  | Nursing Staff | 1 | 2.6 |
|  | Paramedical Staff | 0 | 0 |
| Exposure with surface around patient n=164 | Doctors | 44 | 86.3 |
|  | Nursing Staff | 34 | 87.2 |
|  | Paramedical Staff | 86 | 84.3 |
| Exposure with patient's body fluids via surface n=5 | Doctors | 3 | 5.9 |
|  | Nursing Staff | 1 | 2.6 |
|  | Paramedical Staff | 1 | 1 |

**Table 2:** Type of exposure among the study participants (N=192)

| Type of Exposure | Frequency (% among all HCW) |
| --- | --- |
| **Close contact exposure** | 122 (63.5%) |
| Prolonged face-to-face exposure | 27 (14.1%) |
| Exposure during aerosolizing procedures | 10 (5.2%) |
| Direct Exposure with body fluid | 13(6.8%) |
| **Patient's materials exposure** | 58 (30.2%) |
| Patient's body fluid via materials exposure | 4 (2.1%) |
| **Surface exposure** | 164 (85.4%) |
| Patient's body fluid via surface around patient | 5 (2.6%) |

**Supplementary Table 1** shows various kind of exposure to COVID-19 patient across their profession. Majority of nurses and doctors were exposed in direct contact including face to face and aerosol and direct patient body fluid exposure.

Usage of various PPE during recent contact with COVID-19 positive patient is shown in figure 4. We observed a trend in adherence to PPE with almost all were following the PPE protocol during high-risk procedure including aerosol procedure while less so during direct face to face contact. Almost three-fourth of the HCW were wearing mask, while face shield and glasses were used by only one-fourth during face to face prolong exposure. We also observed that majority of the participants were wearing almost all the PPE while Aerosol Procedure and body fluids exposure.

**Supplementary Table 2:** Adherence to various PPE among doctors, nurses and paramedics during recent contact with COVID-19 patient.

|  | **Any contact**<br>**N=187\*** | **Doctor**<br>**N=50** | **Nurse**<br>**N=38** | **Paramedics**<br>**N=99** |
| --- | --- | --- | --- | --- |
| **Medical/Surgical Mask** | 135 (72.2%) | 44 (88.0%) | 27 (71.1%) | 64 (64.6%) |
| **Respirator** | 152 (81.3%) | 39 (78.0%) | 37 (97.4%) | 76 (76.8%) |
| **Face Shield** | 49 (26.2%) | 14 (28.0%) | 18 (47.4%) | 17 (17.2%) |
| **Gloves** | 112 (59.9%) | 33 (66.0%) | 34 (89.5%) | 45 (45.5%) |
| **Goggles/Glasses** | 59 (31.6%) | 21 (42.0) | 19 (50.0%) | 19 (19.2%) |
| **Gown** | 98 (52.4%) | 27 (54.0%) | 36 (94.7%) | 35 (35.4%) |
| **Coverall** | 44 (23.5%) | 19 (38.0%) | 16 (42.1%) | 9 (9.1%) |
| **Head Cover** | 104 (55.6%) | 27 (54.0%) | 34 (89.5%) | 43 (43.4%) |
| **Shoe Cover** | 98 (52.4%) | 26 (52.0%) | 32 (84.2%) | 40 (40.4%) |

\*5 HCW did not responded to this question

**Supplementary Table 3:.**
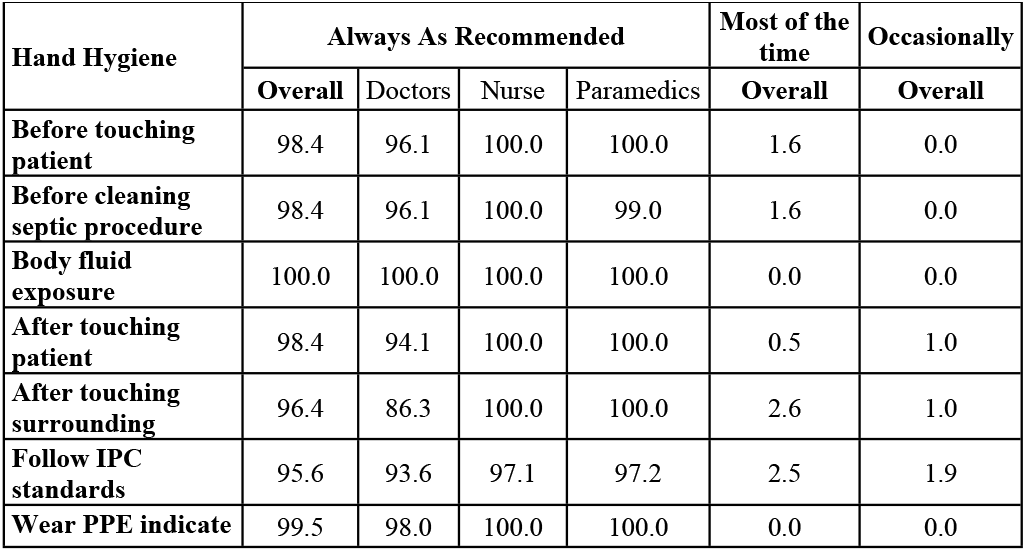
Hand hygine practiced by the healthcare workers.

| **Hand Hygiene** | **Always As Recommended** |  |  |  | **Most of the time** | **Occasionally** |
| --- | --- | --- | --- | --- | --- | --- |
|  | **Overall** | **Doctors** | **Nurse** | **Paramedics** | **Overall** | **Overall** |
| **Before touching patient** | 98.4 | 96.1 | 100.0 | 100.0 | 1.6 | 0.0 |
| **Before cleaning septic procedure** | 98.4 | 96.1 | 100.0 | 99.0 | 1.6 | 0.0 |
| **Body fluid exposure** | 100.0 | 100.0 | 100.0 | 100.0 | 0.0 | 0.0 |
| **After touching patient** | 98.4 | 94.1 | 100.0 | 100.0 | 0.5 | 1.0 |
| **After touching surrounding** | 96.4 | 86.3 | 100.0 | 100.0 | 2.6 | 1.0 |
| **Follow IPC standards** | 95.6 | 93.6 | 97.1 | 97.2 | 2.5 | 1.9 |
| **Wear PPE indicate** | 99.5 | 98.0 | 100.0 | 100.0 | 0.0 | 0.0 |

**Figure 4:**
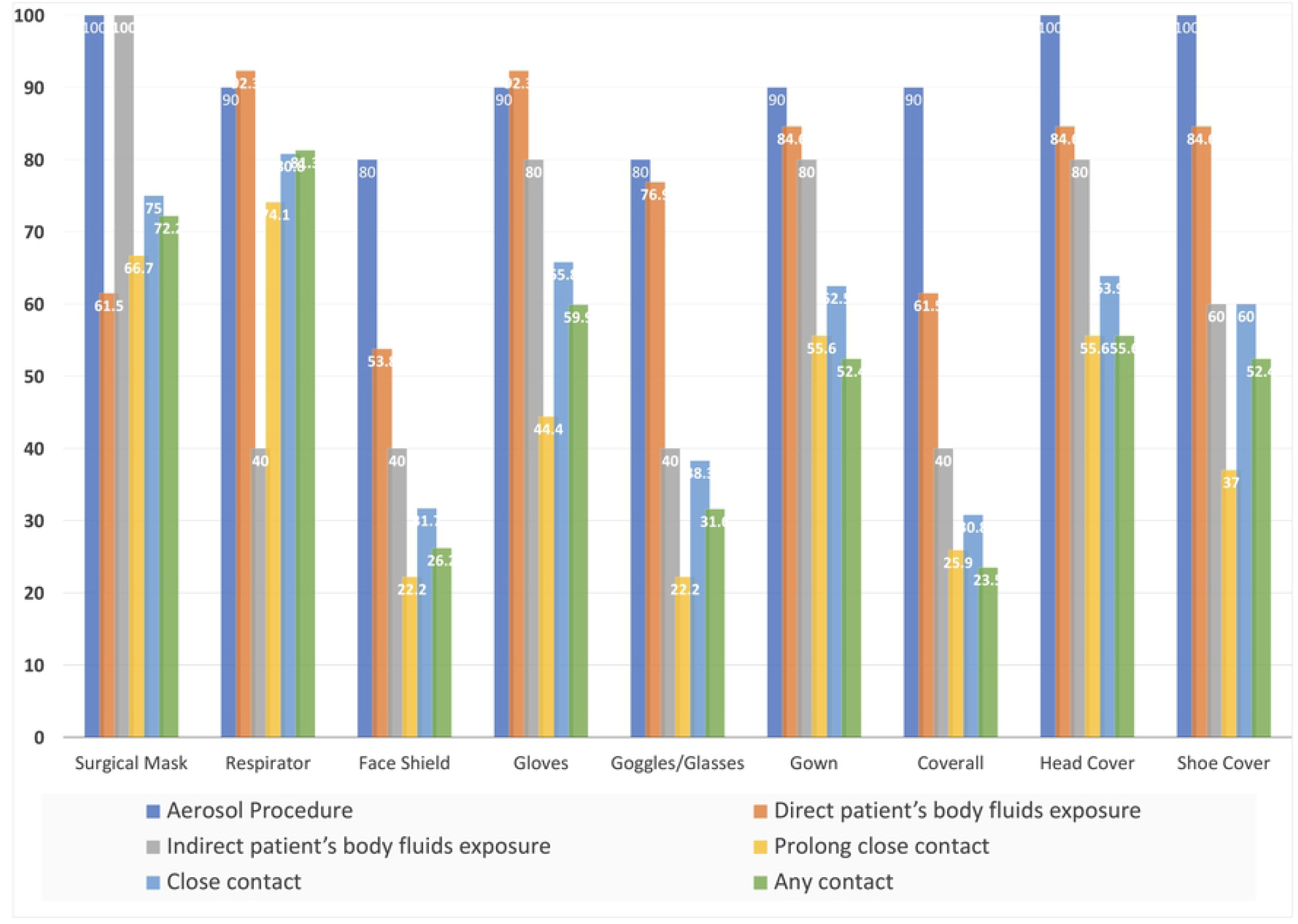
Usage of various PPE by HCW during recent exposure to COVID-19 patient across different type of exposure.

We observed a definite trend of adherence to PPE among various health workers as shown in Supplementary Table 2. The nurses were found to be using most of the PPE during the recent close contact with the COVID-19 case, while paramedics were less adherent, doctors were in between the two in usage of PPE. Hand hygiene practice is also shown in Supplementary Table 3.

**Table 3:** Distribution of Seroconversion and antibody titre increase of HCW at endline.

|  | Seroconversion N= 49 |  | Sero increase n-139 |  |
| --- | --- | --- | --- | --- |
|  | **Frequency** | **% (Secondary infection rate)** | **Frequency** | **%** |
| Doctor | 12 | 63.2% | 25 | 71.4% |
| Nurses | 3 | 42.9% | 14 | 53.8% |
| Paramedics | 3 | 13.0% | 50 | 64.1% |
| Overall | 18 | 36.7% | 89 | 64.0% |

The figure 5 is showing serology status of HCW at baseline, 62% were seropositive. Among all the doctors, 47.1% were seropositive, while 71.6% of paramedical staff were seropositive. In our study only 5.7% had symptoms after the recent contact with the COVID-19 patients admitted in the health care settings, while 7.2% of them had one during their follow up period. The HCW who had symptoms were observed to be less commonly seropositive both at baseline as well as endline. Most common symptoms were headache and fatigue (Supplementary **Table 4)**.

**Supplementary Table 4:** Distribution of Symptom Profile with the serology at baseline (N=192)

|  | **Baseline** |  | **Follow up** |  |
| --- | --- | --- | --- | --- |
|  | Total n=192 | Seropositive<br>n=119<br>(% among<br>seropositive) | Total n=139 | Seropositive<br>n=108<br>(% among<br>seropositive) |
| **Any Symptom** | 11 (5.7%) | 3 (2.5%) | 14 (10.1%) | 5 (4.6%) |
| **Fever** | 0 (0%) | 0 (0%) | 2 (1.4%) | 1 (0.9) |
| **Sore throat** | 2 (1.0%) | 1 (0.8%) | 6 (4.3%) | 3 (2.8) |
| **Cough** | 2 (1.0%) | 1 (0.8%) | 4 (2.9%) | 2 (1.9) |
| **Shortness of**<br>**breath** | 2 (1.0%) | 0 (0%) | 1 (0.7%) | 1 (0.9) |
| **Rhinitis** | 0 (0%) | 0 (0%) | 4 (2.9%) | 1 (0.9) |
| **Chills** | 1 (0.5%) | 1 (0.8%) | 0 (0%) | 0 (0%) |
| **Nausea** | 1 (0.5%) | 0 (0%) | 0 (0%) | 0 (0%) |
| **Vomiting** | 0 (0%) | 0 (0%) | 1 (0.7%) | 0 (0%) |
| **Headache** | 5 (2.6%) | 2 (1.7%) | 2 (1.4%) | 0 (0%) |
| **Rash** | 1 (0.5%) | 0 (0%) | 0 (0%) | 0 (0%) |
| **Muscle aches** | 1 (0.5%) | 0 (0%) | 2 (1.4%) | 1 (0.9) |
| **Joint ache** | 1 (0.5%) | 1 (0.8%) | 0 (0%) | 0 (0%) |
| **Loss of appetite** | 1 (0.5%) | 0 (0%) | 0 (0%) | 0 (0%) |
| **Loss of Sense of taste** | 1 (0.5%) | 0 (0%) | 0 (0%) | 0 (0%) |
| **Fatigue** | 2 (1.0%) | 2 (1.7%) | 4 (2.9%) | 1 (0.9) |
| **Other symptoms** | 1 (0.5%) | 1 (0.8%) | 0 (0%) | 0 (0%) |

**Table 4.** Univariate regression model of variables related to seroprevalence, seroconversion and rise in titre.

| Variable |  | Seroprevalence |  | Seroconversion |  | Rise in titre |  |
| --- | --- | --- | --- | --- | --- | --- | --- |
|  |  | OR (95%CI) | p value | OR (95%CI) | p value | OR (95%CI) | p value |
| **Gender** | Males | 1 |  | 1 |  | 1 |  |
|  | Female | 0.715 (0.398 - 1.283) | 0.26 | 1.551 (0.573 - 4.201) | 0.39 | 0.69 (0.34 - 1.38) | 0.29 |
| **Age groups** | 18-29 | 1 |  | 1 |  | 1 |  |
|  | 30-59 | 2.234 (1.191 - 4.192) | 0.01 | 0.382 (0.118 - 1.238) | 0.11 | 1.14 (0.56 - 2.31) | 0.71 |
|  | 60 and above | 0.00 (0.00 – NaN) | 1.000 | High (0.00 – NaN) | 1.000 | HIGH (0.000- NaN) | 1.00 |
| **Profession** | Doctor | 0.353 (0.176- 0.710) | 0.003 | 13.043 (3.386 - 50.246) | <0.001 | 1.40 (0.59 - 3.33) | 0.45 |
|  | Nurse | 0.514 (0.239- 1.105) | 0.09 | 3.261 (0.616 - 17.272) | 0.17 | 0.65 (0.27 - 1.61) | 0.35 |
|  | Paramedic | 1 |  | 1 |  | 1 |  |
| **Work place** | High risk | 0.569 (0.316 - 1.025) | 0.06 | 1.136 (0.419 - 3.081) | 0.80 | 0.98 (0.49 - 1.98) | 0.96 |
|  | Low risk | 1 |  | 1 |  | 1 |  |
| **Smoking** | Yes | 0.840 (0.352- 2.004) | 0.69 | 0.00 (0.00 – NaN) | 0.99 | 0.35 (0.13 - 0.94) | 0.04 |
|  | No | 1 |  | 1 |  | 1 |  |
| **Comorbidities** | Yes | 0.139 (0.029 - 0.674) | 0.01 | 0.00 (0.00 – NaN) | 0.99 | 0.08 (0.01 - 0.71) | **0.02** |
|  | No | 1 |  | 1 |  | 1 |  |
| **COVID-19 symptoms** | Yes | 0.210 (0.054- 0.820) | 0.025 | 0.489 (0.113 – 2.11) | 0.337 | 0.188 (0.056 – 0.637) | 0.007 |
|  | No | 1 |  | 1 |  | 1 |  |
| **Vaccination** | Not Vaccinated | 1 |  | 1 |  | 1 |  |
|  | Partially Vaccinated | 3.303 (1.256- 8.685) | 0.02 | 4.345 (1.070 - 17.647) | 0.04 | 7.87 (2.18 - 28.40) | 0.002 |
|  | Vaccinated | 2.428 (1.118- 5.271) | 0.03 | 6.083 (1.729 - 21.399) | 0.005 | 3.59 (1.46 - 8.87) | 0.006 |
| **IPC Training** | Not received | 1 |  | 1 |  | 1 |  |
|  | Less than 2 hr | 0.485 (0.184- 1.284) | 0.15 | 1.689 (0.358 - 7.965) | 0.51 | 1.90 (0.73 - 4.97) | 0.19 |
|  | More than 2 hr | 0.250 (0.072 - 0.864) | 0.03 | 0.00 (0.00 – NaN) | 0.99 | 0.07 (0.01 - 0.63) | 0.02 |

\*\*NaN is Not a Number, unable to divide by 0.

**Figure 5:**
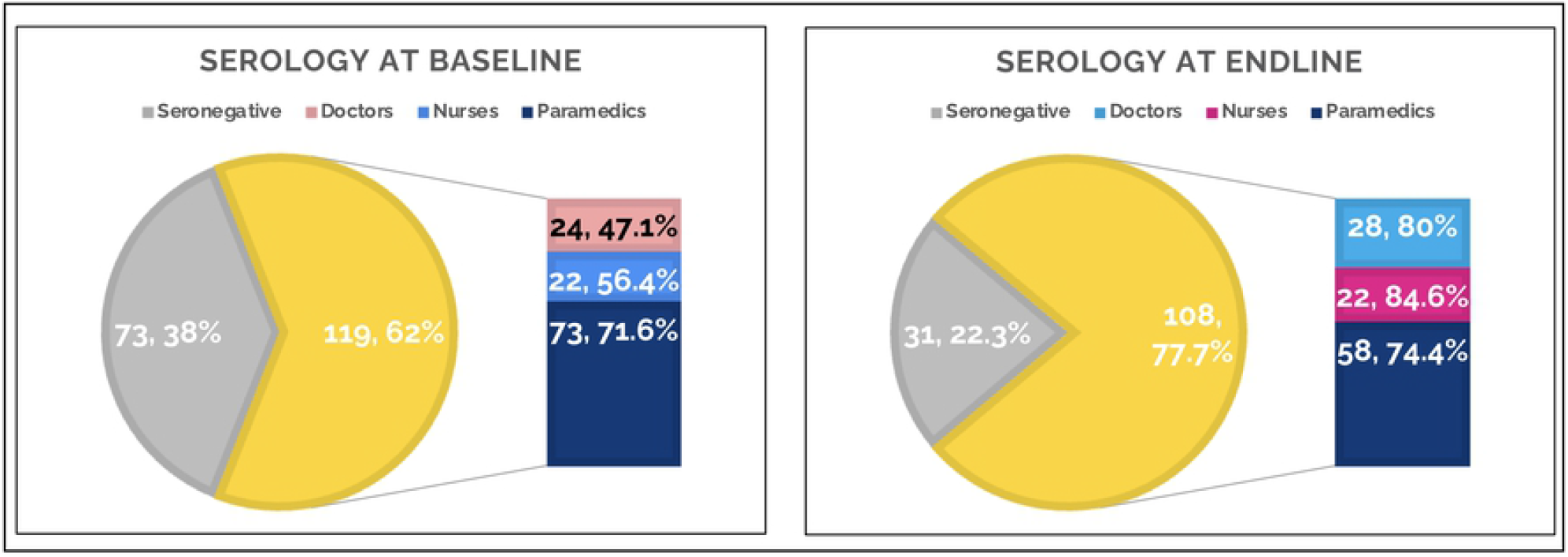
Distribution of serology status against COVID19 of HCW at baseline and endline.

Figure 6 shows seroconversion rate and serial rise in antibody titre between baseline and endline among HCW. We had observed that more than one third (36.7%) of the HCW became positive for the antibody against COVID-19 at endline who were negative at baseline. In term of rise in titre, it was observed in 64.0% of HCW. The seroconversion rate was 63.2% among doctors, 42.9% in nurses and 13.0% in paramedical staff. Doctors antibody titre was observed to increase maximum (71.4%) while in nurses it was seen in 53.8%. (Table 3)

**Supplementary Table 5:**
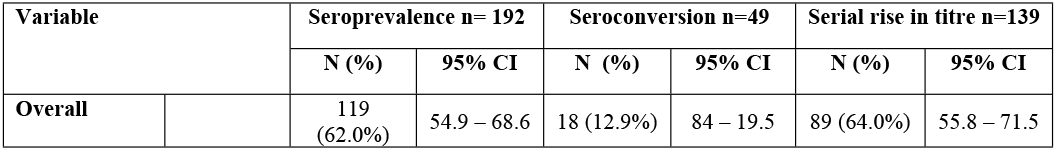
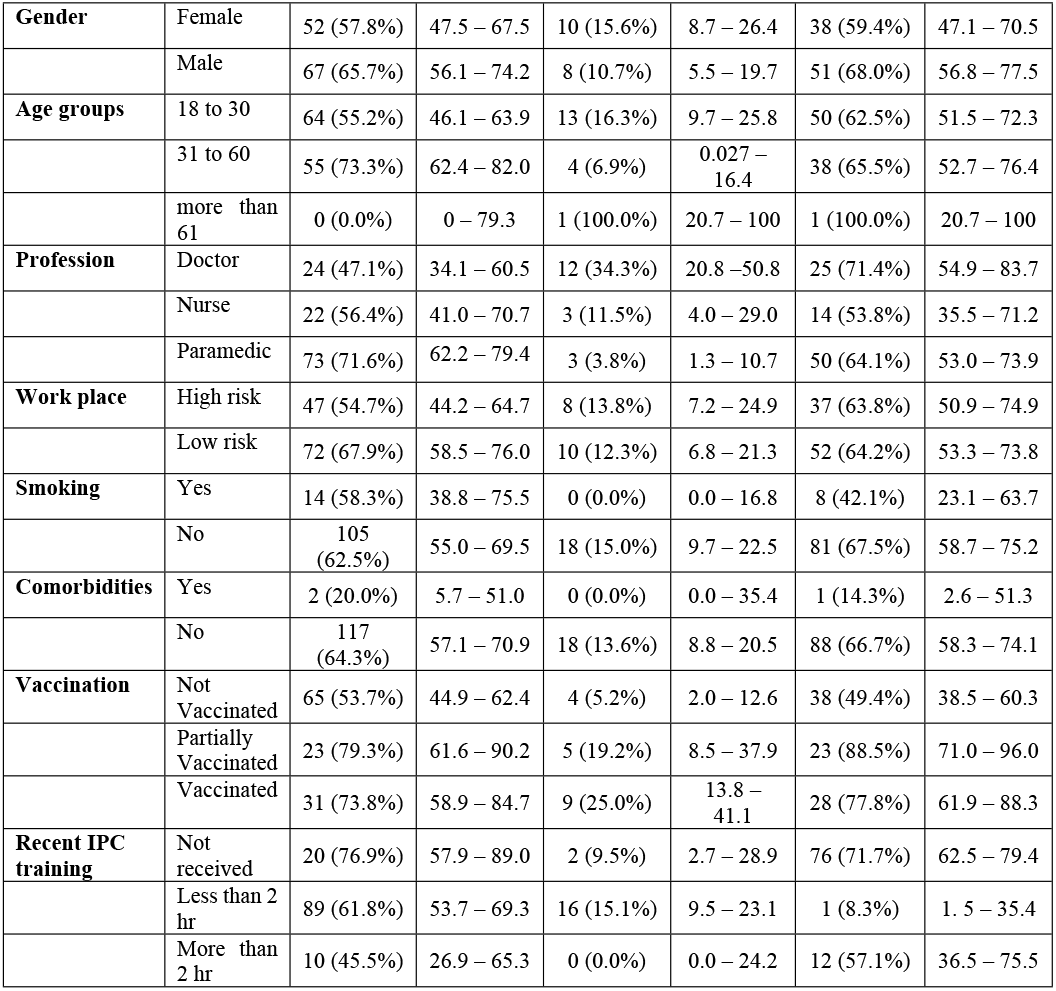
Overall seroprevalence and seroconversion among healthcare workers.

| Variable |  | Seroprevalence n= 192 |  | Seroconversion n=49 |  | Serial rise in titre n=139 |  |
| --- | --- | --- | --- | --- | --- | --- | --- |
|  |  | **N (%)** | **95% CI** | **N (%)** | **95% CI** | **N (%)** | **95% CI** |
| **Overall** |  | 119 (62.0%) | 54.9 – 68.6 | 18 (12.9%) | 84 – 19.5 | 89 (64.0%) | 55.8 – 71.5 |
| **Gender** | Female | 52 (57.8%) | 47.5 – 67.5 | 10 (15.6%) | 8.7 – 26.4 | 38 (59.4%) | 47.1 – 70.5 |
|  | Male | 67 (65.7%) | 56.1 – 74.2 | 8 (10.7%) | 5.5 – 19.7 | 51 (68.0%) | 56.8 – 77.5 |
| **Age groups** | 18 to 30 | 64 (55.2%) | 46.1 – 63.9 | 13 (16.3%) | 9.7 – 25.8 | 50 (62.5%) | 51.5 – 72.3 |
|  | 31 to 60 | 55 (73.3%) | 62.4 – 82.0 | 4 (6.9%) | 0.027 – 16.4 | 38 (65.5%) | 52.7 – 76.4 |
|  | more than 61 | 0 (0.0%) | 0 – 79.3 | 1 (100.0%) | 20.7 – 100 | 1 (100.0%) | 20.7 – 100 |
| **Profession** | Doctor | 24 (47.1%) | 34.1 – 60.5 | 12 (34.3%) | 20.8 – 50.8 | 25 (71.4%) | 54.9 – 83.7 |
|  | Nurse | 22 (56.4%) | 41.0 – 70.7 | 3 (11.5%) | 4.0 – 29.0 | 14 (53.8%) | 35.5 – 71.2 |
|  | Paramedic | 73 (71.6%) | 62.2 – 79.4 | 3 (3.8%) | 1.3 – 10.7 | 50 (64.1%) | 53.0 – 73.9 |
| **Work place** | High risk | 47 (54.7%) | 44.2 – 64.7 | 8 (13.8%) | 7.2 – 24.9 | 37 (63.8%) | 50.9 – 74.9 |
|  | Low risk | 72 (67.9%) | 58.5 – 76.0 | 10 (12.3%) | 6.8 – 21.3 | 52 (64.2%) | 53.3 – 73.8 |
| **Smoking** | Yes | 14 (58.3%) | 38.8 – 75.5 | 0 (0.0%) | 0.0 – 16.8 | 8 (42.1%) | 23.1 – 63.7 |
|  | No | 105 (62.5%) | 55.0 – 69.5 | 18 (15.0%) | 9.7 – 22.5 | 81 (67.5%) | 58.7 – 75.2 |
| **Comorbidities** | Yes | 2 (20.0%) | 5.7 – 51.0 | 0 (0.0%) | 0.0 – 35.4 | 1 (14.3%) | 2.6 – 51.3 |
|  | No | 117 (64.3%) | 57.1 – 70.9 | 18 (13.6%) | 8.8 – 20.5 | 88 (66.7%) | 58.3 – 74.1 |
| **Vaccination** | Not Vaccinated | 65 (53.7%) | 44.9 – 62.4 | 4 (5.2%) | 2.0 – 12.6 | 38 (49.4%) | 38.5 – 60.3 |
|  | Partially Vaccinated | 23 (79.3%) | 61.6 – 90.2 | 5 (19.2%) | 8.5 – 37.9 | 23 (88.5%) | 71.0 – 96.0 |
|  | Vaccinated | 31 (73.8%) | 58.9 – 84.7 | 9 (25.0%) | 13.8 – 41.1 | 28 (77.8%) | 61.9 – 88.3 |
| **Recent IPC training** | Not received | 20 (76.9%) | 57.9 – 89.0 | 2 (9.5%) | 2.7 – 28.9 | 76 (71.7%) | 62.5 – 79.4 |
|  | Less than 2 hr | 89 (61.8%) | 53.7 – 69.3 | 16 (15.1%) | 9.5 – 23.1 | 1 (8.3%) | 1.5 – 35.4 |
|  | More than 2 hr | 10 (45.5%) | 26.9 – 65.3 | 0 (0.0%) | 0.0 – 24.2 | 12 (57.1%) | 36.5 – 75.5 |

**Figure 6:**
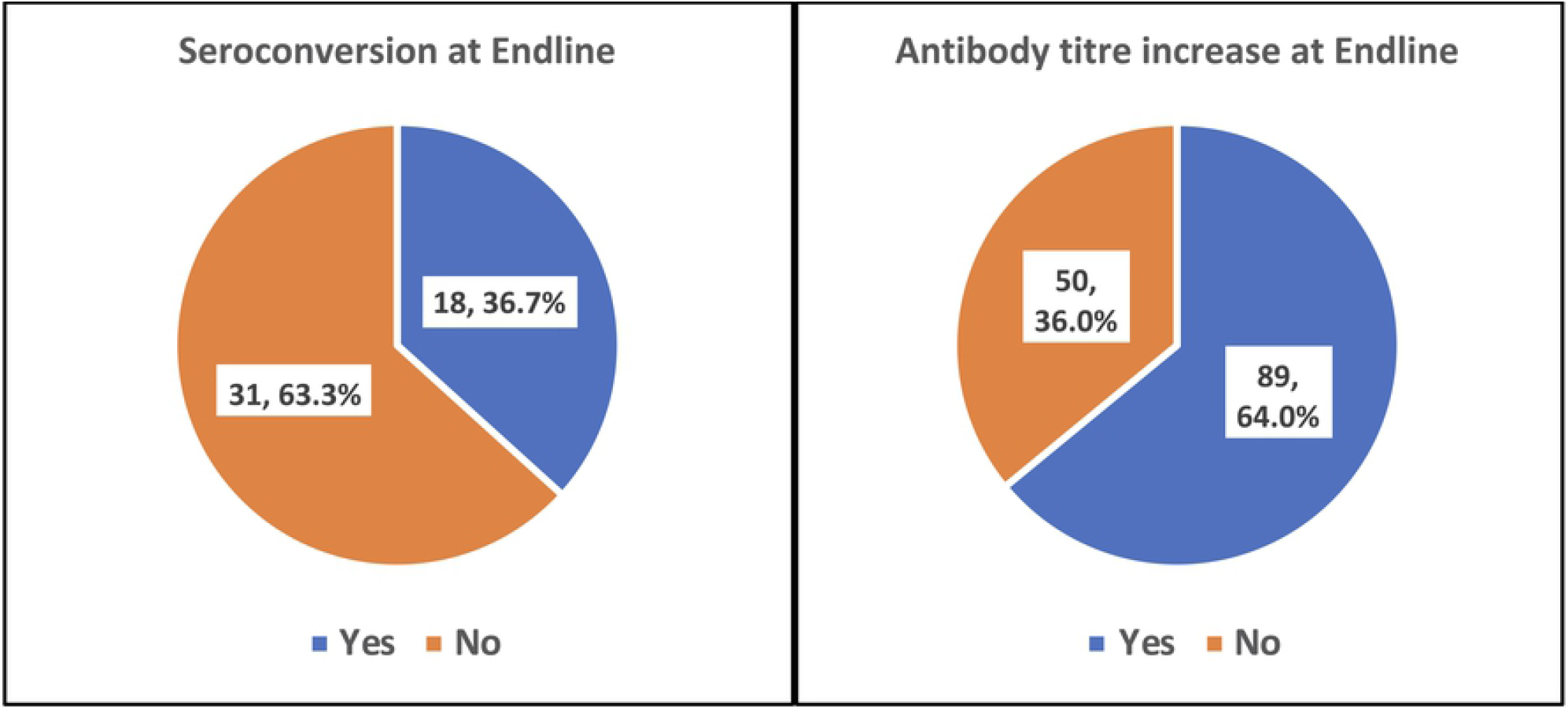
Seroconversion rate and antibody titre increase of HCW at endline.

Supplementary Table 5 shows prevalence of seropositivity and seroconversion of study population along with their 95% confidence intervals. We observed seropositivity of 62% (95% CI: 54.9 – 68.6) among our recruited HCW working in the tertiary health centre in New Delhi. We also observed high seropositivity among various factors including higher age group (73.3%), Paramedic staff (71.6%), working in low-risk area (67.9%), vaccinated (73.8%) and not received Infection prevention control training (76.9%).

We also assessed the risk of seropositivity, seroconversion and rise in titre as highlighted in table 4. The seropositivity was significantly and negatively associated with doctor as profession [OR:0.353, CI:0.176-0.710], COVID-19 symptoms [OR:0.210, CI:0.054-0.820], comorbidities [OR:0.139, CI: 0.029 - 0.674], recent IPC Training [OR:0.250, CI:0.072 - 0.864], while positively associated with middle age [OR 2.234, CI:1.191 - 4.192], partially [OR:3.303,CI: 1.256-8.685], as well as fully Vaccinated for COVID-19 [OR:2.428, CI:1.118-5.271]. The seroconversion was observed to be significantly and positively associated with doctors [OR:13.043; CI:3.386 – 50.246] and partially [OR: 4.345, CI: 1.070 - 17.647], as well as fully Vaccinated for COVID-19 [OR: 6.083, CI: 1.729 - 21.399]. The serial rise in trite was significantly and negatively associated with serial rise in titre of antibodies with history of symptoms [OR: 0.188; CI: 0.056 – 0.637], smokers[OR: 0.35, 95%CI: 0.13 - 0.94], HCW with comorbidities [OR:0.08,95CI: 0.01 - 0.71], recent full IPC Training [OR:0.07, CI:0.01 - 0.63], while positively associated with partially [OR: 7.87, 95CI: 2.18 - 28.40)], as well as fully vaccinated for COVID-19 [OR: 3.59, 95CI: 1.46 - 8.87].

In our study we did not observe any association of type of exposure type as well as adherence of PPE and IPC practices with both seroconversion and serial rise in titre of antibodies against the COVID-19 after 3-4 weeks. (Table 5 & Table 6)

**Supplementary Table 6:**
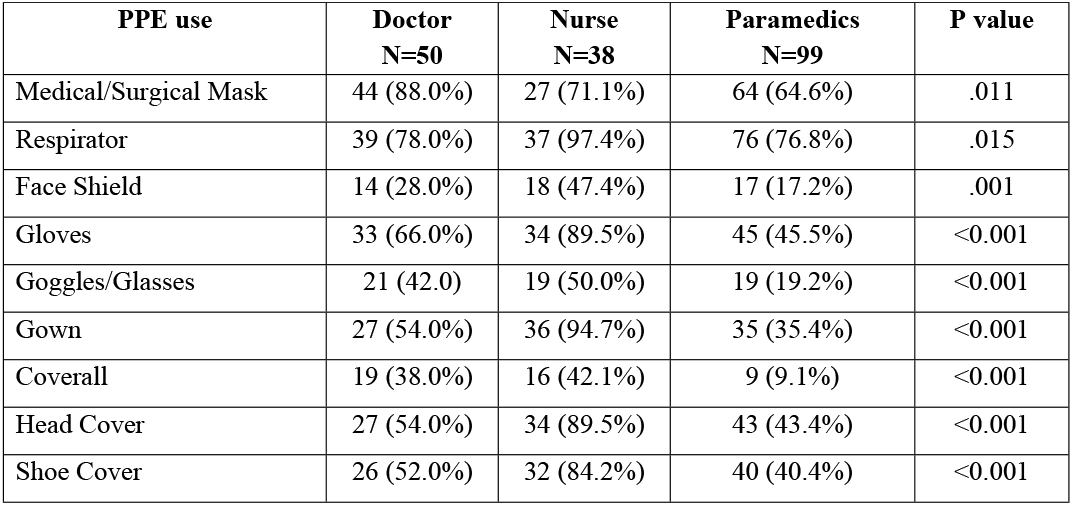
Association of Adherence to various PPE during recent prolong close contact with COVID-19 patient with their profession.

| **PPE use** | **Doctor**<br>**N=50** | **Nurse**<br>**N=38** | **Paramedics**<br>**N=99** | **P value** |
| --- | --- | --- | --- | --- |
| Medical/Surgical Mask | 44 (88.0%) | 27 (71.1%) | 64 (64.6%) | .011 |
| Respirator | 39 (78.0%) | 37 (97.4%) | 76 (76.8%) | .015 |
| Face Shield | 14 (28.0%) | 18 (47.4%) | 17 (17.2%) | .001 |
| Gloves | 33 (66.0%) | 34 (89.5%) | 45 (45.5%) | <0.001 |
| Goggles/Glasses | 21 (42.0) | 19 (50.0%) | 19 (19.2%) | <0.001 |
| Gown | 27 (54.0%) | 36 (94.7%) | 35 (35.4%) | <0.001 |
| Coverall | 19 (38.0%) | 16 (42.1%) | 9 (9.1%) | <0.001 |
| Head Cover | 27 (54.0%) | 34 (89.5%) | 43 (43.4%) | <0.001 |
| Shoe Cover | 26 (52.0%) | 32 (84.2%) | 40 (40.4%) | <0.001 |

**Table 5:**
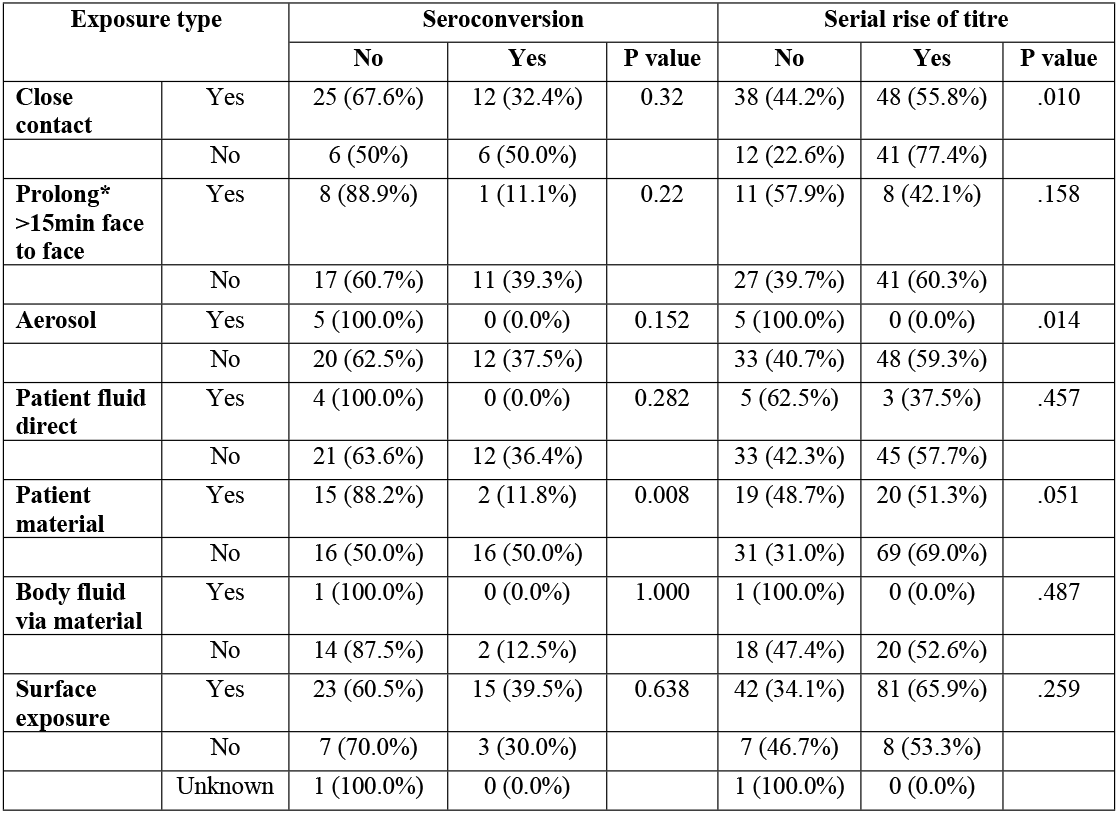
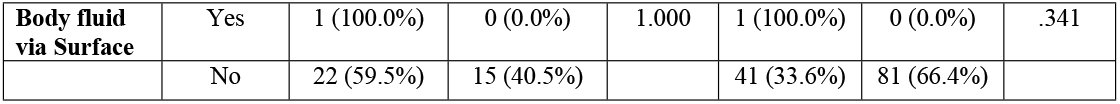
Exposure type along with IPC measures among the study participants and seroconversion.

**Table 6:** Adherence of PPE and Seroconversion and Serial rise of titre.

| PPE wore |  | Seroconversion |  |  | Serial rise of titre |  |  |
| --- | --- | --- | --- | --- | --- | --- | --- |
|  |  | No | Yes | P value | No | Yes | P value |
| **Surgical Mask** | Yes | 24 (64.9%) | 13 (35.1%) | 0.738 | 35 (36.5%) | 61 (63.5%) | 0.989 |
|  | No | 7 (58.3%) | 5 (41.7%) |  | 15 (36.6%) | 26 (63.4%) |  |
| **Respirator** | Yes | 25 (62.5%) | 15 (37.5%) | 1.000 | 40 (35.7%) | 72 (64.3%) | 0.819 |
|  | No | 6 (66.7%) | 3 (33.3%) |  | 10 (40.0%) | 15 (60.0%) |  |
| **Face Shield** | Yes | 7 (87.5%) | 1 (12.5%) | 0.229 | 12 (33.3%) | 24 (66.7%) | 0.691 |
|  | No | 24 (58.5%) | 17 (41.5%) |  | 38 (37.6%) | 63 (62.4%) |  |
| **Gloves** | Yes | 18 (64.3%) | 10 (35.7%) | 0.864 | 29 (35.8%) | 52 (64.2%) | 0.858 |
|  | No | 13 (61.9%) | 8 (38.1%) |  | 21 (37.5%) | 35 (62.5%) |  |
| **Goggles/ Glasses** | Yes | 12 (66.7%) | 6 (33.3%) | 0.767 | 16 (38.1%) | 26 (61.9%) | 0.848 |
|  | No | 19 (61.3%) | 12 (38.7%) |  | 34 (35.8%) | 61 (64.2%) |  |
| **Gown** | Yes | 15 (65.2%) | 8 (34.8%) | 0.790 | 26 (37.1%) | 44 (62.9%) | 0.872 |
|  | No | 16 (61.5%) | 10 (38.5%) |  | 24 (35.8%) | 43 (64.2%) |  |
| **Coverall** | Yes | 5 (62.5%) | 3 (37.5%) | 0.961 | 10 (34.5%) | 19 (65.5%) | 0.800 |
|  | No | 26 (63.4%) | 15 (36.6%) |  | 40 (37.0%) | 68 (63.0%) |  |
| **Head Cover** | Yes | 17 (73.9%) | 6 (26.1%) | 0.146 | 27 (35.5%) | 49 (64.5%) | 0.792 |
|  | No | 14 (53.8%) | 12 (46.2%) |  | 23 (37.7%) | 38 (62.3%) |  |
| **Shoe Cover** | Yes | 17 (73.9%) | 6 (26.1%) | 0.146 | 25 (35.2%) | 46 (64.8%) | 0.746 |
|  | No | 14 (53.8%) | 12 (46.2%) |  | 25 (37.9%) | 41 (62.1%) |  |
| **Remove gloves after contact** | yes | 18 (62.1%) | 11 (37.9%) | 0.834 | 33 (35.1%) | 61 (64.9%) | .759 |
|  | no | 13 (65.0%) | 7 (35.0%) |  | 17 (37.8%) | 28 (62.2%) |  |
| **Hand hygiene performed** | Adequate | 28 (63.6%) | 16 (36.4%) | 1.00 | 46 (34.6%) | 87 (65.4%) | .188 |
|  | Not adequate | 3 (60%) | 2 (40%) |  | 4 (66.7%) | 2 (33.3%) |  |

As shown in Supplementary Table 6, the nurses were found to be using most of the PPE during the recent close contact with the COVID-19 case, while paramedics were less adherent, doctors were in between the two in usage of PPE. This association was found to be statistically significant in all the type of PPE.

## DISCUSSION

We did this study with the objective of assessing human to human transmission by measuring the seroconversion of COVID-19 among recently exposed healthcare workers and to evaluate the effectiveness of infection prevention and control measures. This was done in New Delhi, the city which was hard hit during first and second wave of COVID-19 pandemic in India. In this we collected information about the exposure to COVID-19 and their risk factors, along with the serological testing at baseline and after 3-4 week, along with daily symptoms by symptoms diary. Our results show that the seropositivity was 62.0%. We found that seropositivity significantly and negatively associated with doctor as profession, having symptoms, comorbidities, recent IPC training while positively associated with partially as well as fully vaccinated for COVID-19.

This research confirms the previous studies of higher seroprevalence of antibodies against the COVID-19 among health care workers. We are of the opinion that these may be due to higher risk of transmission as well as the ongoing immunization program against COVID-19. We also observed seroconversion in 36.7% HCW while 64.0% had serial rise in titre of antibodies during our follow-up period. The seroconversion was higher in doctors, and nurses (63.2% and 42.9% respectively) in comparison to paramedics staff (13.0%). The seroconversion was positively associated with doctor as profession and with partially, as well as fully vaccinated for COVID-19. None of the HCW who were smokers and with any comorbidity had seroconversion. We observe a negative and significant relationship of serial rise in titre of antibodies with recent influenza like illness (ILI), smoker, HCW with comorbidities, and the recent full IPC training, while positively associated with partially, as well as full vaccination for COVID-19. Adherences to the infection prevention measure adopted by the HCW during the recent contact with COVID-19 patients was not found to be significantly associated with seroconversion or serial rise in titre.

Among the HCW we enrolled, more than half were paramedical staff and males and belonging to young age group. Studies done among HCW have varied finding, with majority supporting the similar health care worker profiles like our study. (12). With regard to comorbidity, our finding are in contrast to the healthcare setting in Chile, where comorbidities were seen in almost half while smoking in a quarter of them(13). Only a few (5.7%) of the HCW had symptoms at the baseline, and majority of those who had symptoms were seropositive. This is supported by countrywide sero-survey which observed majority of the seropositive participants were not having any kind of symptoms (14). It has been found in studies done across the continent that majority of the COVID-19 patients are asymptomatic, while only a few require hospitalization (15). Our findings is contrast to the observations of researcher from Spain, where they found higher proportion of HCWs with comorbidities and COVID-19-compatible symptoms in the previous months(16).

We also documented incubation period in our study which was 7.9 ± 6.8 days (median: 7.5; IQR:1.0 – 15.0). The CDC reports incubation period of COVID-19 to extending up to 14 days with median of 4-5 days(17).

We observed that all the doctors and nurses and almost all paramedical staff were wearing some kind of PPE when they were exposed to COVID-19 patient. This was expected because the of mandatory PPE policy adopted by the hospital where this study was conducted. The PPE adherence was seen high risk procedure, but less in low risk activity specially by the paramedics, who were even found to be significantly non adherent in usage of PPE. We did not observe any significant association usage of individual PPE with seroconversion as well as increase in titre. Studies has shown appropriate use of PPE to be the most critical defence against COVID-19 infection among HCW (18)(19)(20). This could not be observed in our study may be due to the very high adherence of appropriate PPE in high-risk places, leading to non-significant relation as well as due to the confounding by vaccination of the HCW.

While two-third doctors and a majority of nurses (82%) were performing appropriate hand hygiene practices while in contact with COVID-19 patient, only half of the paramedical staff were following the same. This could be due to the fact that doctors and nurses are involved in direct patient care, while paramedical staff, who are not directly involve in patient care may consider themselves at lesser risk. This may be further due to the differences in information available about importance of adherence to IPC practices. Unlike our study, an international study observed higher knowledge and practice of PPE in non-physicians in comparison to physicians (21). Similar to ours, a study also observed that resident doctors and paramedic reported lower adherence to PPE (22) This along with not performing hand hygiene practices could be one of the major potential for exposure to the COVID-19. Previous studies have also stressed on IPC practices among various categories of HCW(23)(24).

In this study HCW were exposed to close contact exposure, face to face prolong exposure, aerosol generating procedure, exposure with patients material, patient’s body fluid, and environment surface exposure. All of these exposures made them a high-risk group for contracting COVID-19. There are many studies which demonstrate HCW being infected due to these procedures(25)(26). Inspite of this, we did not observe any association of type of exposure with seroconversion except among those exposure to patient material. This might be due to good adherence to PPE and IPC measures in our hospital setting, as well as due to confounding by concurrent vaccination program.

COVID-19 Vaccination was observed to be the most important factors associated/confounded with the seropositivity and seroconversion of the HCW in this study. This was not considered in the initial planning of this study, and we have to make a protocol deviation on 1^st^ March 2021, enrolling HCW who were vaccinated, with or without seropositive at baseline. About two third (63%) of the HCW were not vaccinated; nurses and paramedics were higher in proportion of unvaccinated. Among the vaccinated group of HCW, doctors were a majority. These proportion are observed among higher side in comparison national figure among general population as well as overall health care worker. (27) Studies from other countries have varied finding with US HCW have higher vaccination coverage while developing countries rates are far behind. US and other developed nation have better vaccination rate, however in developing countries like India (6.2%), Brazil(16%) and Bangladesh(2.6%) has comparatively low vaccination coverage(28). Using unvaccinated as a reference population we found that vaccination to be strongly and positively associated with seropositivity [OR:2.428, CI:1.118-5.271], seroconversion [OR: 6.08, CI: 1.729 - 21.40] as well as serial rise in titre [OR: 3.59, 95CI: 1.46 - 8.87]. These findings were expected since COVID-19 vaccines are given to produce antibodies by the immune system(29). This was true in our study where we observed the strongest association of vaccination with seroconversion.

The seropositivity at baseline was 62% while at endline was observed to be 77.7%. Our study findings are supported by a study where positive IgG response was observed in 48.4 % and 77.8% at baseline and follow up respectively(30). The latest sero-survey in India, observed low seropositivity among general population. A sero-survey done in Delhi has also observed low seropositive rate in contrast to our study(31). One Indian study found doctors have higher risk of seropositivity than other HCWs which is similar to our study finding (14). Globally the seroprevalence among HCW varied from 3% in Finland to 55.9% in Brazil(32). It was also found that seropositivity was higher in HCWs involved in COVID-19 patient management(33), although a study from Chile found no statistical difference in seropositivity among HCW involved in direct clinical care of patients with COVID-19 and those working in low risk areas(34). We are in an opinion that high seroprevalence in our study papulation is due to the high vaccination rates, which was also one of the strongest risk factors observed in our study.

We also observed various factors associated with seropositivity at baseline. It was observed that doctor as profession [OR:0.353, CI:0.176-0.710], having symptoms [OR:0.210, CI:0.054-0.820], comorbidities [OR:0.139, CI: 0.029 - 0.674], recent IPC Training [OR:0.250, CI:0.072 - 0.864], partially vaccinated [OR:3.303, CI: 1.256-8.685], as well as fully vaccinated for COVID-19 [OR:2.428, CI:1.118-5.271] had significant risk factors for seropositivity. Age and gender of the HCW, usage of PPE and adherence to IPC did not had significant association of seropositivity in our study, while those who had any symptom had lower risk of seroconversion. These are in contrast to study from similar settings in New Delhi observed seropositivity to be associated with male sex (31), while one from Germany observed use of PPE to be protective (35). A study from Spain reported high odds of seropositive among those having any COVID-19 symptom in the previous months, although found no association with profession, working in high risk unit, close contact with a COVID-19 case, comorbidities and sex, partially supporting our findings(16). Higher seroprevalence in paramedical staff in comparison to doctors (71.6% vs 47.1%) as well as those working in low-risk area in comparison to high-risk area (67.9% vs 54.7%). This could be explained by strict adherences to IPC measure and higher use of PPE as seen among allied health workers in our study, who are more often responsible for managing patients in high-risk setting. The health care setting where this study took place, had a dedicated IPC committee, COVID surveillance Unit and proper PPE mandate, leading to better adherence of IPC measures and higher availability of PPEs in comparison to other hospitals and effective contact tracing.

The seroconversion was documented to be 36.7 % among HCW, was 63.2% in doctors, 42.9% in nurses and 13.0% in paramedics staff. We also observed that 64.0% had increase in titre of antibodies during our follow-up period. Studies done across the globe have varied rates of seroconversion. In a large prospective study in UK it was 0.77% (36), 5.4% in Italy(37), 24% in Chile (13) while 44% from Paediatric Dialysis Unit in USA (38). Seroconversion among HCW for H1N1 in 2009 was also documented as 6.5%(39). This variation could be due to different settings and study period. A study from Germany observed rise in titre among 72% of HCW during the follow-up period (35) We are in an opinion that high seroconversion in our study papulation is due to the concurrent vaccination program - which was also one of the strongest risk factors observed in our study - rather than secondary infection from COVID-19 case to which HCW was exposed. Seroconversion was positively associated with doctor as profession [OR: 13.04, CI: 3.39 - 50.25] and with partially [OR: 4.35, CI: 1.070 - 17.647], as well as fully vaccinated for COVID-19 [OR: 6.08, CI: 1.729 - 21.40]. We also observed a negative and significant relationship of serial rise in titre of antibodies with symptoms [OR:0.17, 0.13 - 0.94], smokers [OR: 0.35, 95%CI: 0.13 - 0.94], comorbidities [OR:0.08,95CI: 0.01 - 0.71], recent IPC Training [OR:0.07, CI:0.01 - 0.63], while positively associated with partially [OR: 7.87, 95CI: 2.18 - 28.40)], as well as fully vaccinated for COVID-19 [OR: 3.59, 95CI: 1.46 - 8.87]. None of the HCW who were smokers, with any comorbidity as well as those who had attended adequate IPC training had seroconversion. We observed higher seroconversion among females, higher age group and those working in high-risk setting, but did not reach the level of significance (p>0.05). Our study findings are supported by the various studies done different part of the world(13) (40). Negative association of smoking and seroconversion found in our study is also supported by study from Chile, where they found smokers showing lower seroconversion(13). Unlike our study, researchers in Italy have observed symptoms to be positively associated (37), while more nurses in comparison to allied physicians were significantly more associated with seroconversion for H1N1 in the past (39).

## LIMITATIONS

Inspite of best of our effort, we had a limitation due to concurrent vaccination drive among HCW which confounded our study and thus prevented us in understanding the development of secondary infection among HCW. This could be solved by confirming the infection with RT-PCR testing, but doing it after every contact of HCW with COVID-19 patients would be non-practical and unethical during the time, our health care system was overwhelmed with requirement of COVID testing. We also had a higher attrition then expected, which could bring bias, although the profile of responders and non-responders did not vary significantly.

## CONCLUSION and RECOMMENDATIONS

Our study observed higher seroprevalence among HCW and its associated with vaccination for COVID-19. The seropositivity was high among paramedical staff but more doctors were seroconverted and increased titre after the exposure to COVID-19 patient. Doctors as well as vaccinated HCW were found to be highly associated with seroconversion and protective against the COVID-19. Hence, it is strongly recommended to increase the vaccination coverage for all cadre of HCWs.

This confounding of infection with vaccination may be curtailed using anti N antibodies serology, in future research. It was also observed that PPE, hand hygiene and IPC measures in the facility practiced and are protective. However, these are better followed by nurses and doctors than the paramedical staff. Therefore, imparting frequent IPC trainings to the paramedical staff is vital in preventing COVID-19. We are in opinion that it would be appropriate to regularly test all healthcare workers for COVID-19, using both PCR and serological assays, irrespective of exposure or symptom history so as to protect this workforce.

## Data Availability

The research data are available at public repository Zenodo. https://doi.org/10.5281/zenodo.5703338 https://zenodo.org/record/5703338#.YhqZRi8RpQI

## Acknowledgments

This study was supported by grants from the World Health Organization under UNITY studies. We are thankful to hospital admiration of HIMSR, New Delhi including Dr. G.N Qazi, CEO, and Dr. Ajaz Mustafa, MS. We also would like to sincerely thank all the study participants for their time and cooperation in completing this project.

## Contributors

MD, AS, FI, YA, MA and MP conceived the study and supervised data collection. AS, FI, YA, N, SN and VJ collected the data. YA, VK, and AR conceived the statistical analysis plan. YA, and VK cleaned data. MD, AS, FI, YA, MA, VS, AR and MP drafted the manuscript and figures. All the authors contributed in writing of the manuscript

## Data sharing

Data sharing requests should be directed to the corresponding author.

## REFERENCES

1. Decaro N. Alphacoronavirus‡. In: Tidona C, Darai G, editors. The Springer Index of Viruses [Internet]. New York, NY: Springer; 2011 [cited 2021 Jul 19]. p. 371–83. Available from: https://doi.org/10.1007/978-0-387-95919-1_56

2. Park HY, Lee EJ, Ryu YW, Kim Y, Kim H, Lee H, et al. Epidemiological investigation of MERS-CoV spread in a single hospital in South Korea, May to June 2015*. Eurosurveillance. 2015 Jun 25;20(25):21169.

3. Fagbo SF, Skakni L, Chu DKW, Garbati MA, Joseph M, Peiris M, et al. Molecular Epidemiology of Hospital Outbreak of Middle East Respiratory Syndrome, Riyadh, Saudi Arabia, 2014. Emerg Infect Dis. 2015 Nov;21(11):1981–8.

4. WHO Coronavirus (COVID-19) Dashboard [Internet]. [cited 2022 Feb 26]. Available from: https://covid19.who.int

5. India’s Case Fatality Rate (CFR) falls below 1.5% [Internet]. [cited 2022 Feb 01]. Available from: https://pib.gov.in/Pressreleaseshare.aspx?PRID=1669006

6. Liu C-Y, Yang Y, Zhang X-M, Xu X, Dou Q-L, Zhang W-W, et al. The prevalence and influencing factors in anxiety in medical workers fighting COVID-19 in China: a cross-sectional survey. Epidemiol Infect. 148:e98.

7. Bandyopadhyay S, Baticulon RE, Kadhum M, Alser M, Ojuka DK, Badereddin Y, et al. Infection and mortality of healthcare workers worldwide from COVID-19: a systematic review. BMJ Glob Health. 2020 Dec 1;5(12):e003097.

8. COVID-19: Nearly 25% Of Delhi’s Population Have Developed Antibodies For COVID-19 [Internet]. Latest Asian, Middle-East, EurAsian, Indian News. 2020 [cited 2021 Sep 7]. Available from: https://eurasiantimes.com/indias-first-sero-surveillance-in-delhi-reveals-25-of-population-infected-with-covid-19/

9. Chen Y, Tong X, Wang J, Huang W, Yin S, Huang R, et al. High SARS-CoV-2 antibody prevalence among healthcare workers exposed to COVID-19 patients. J Infect. 2020 Sep 1;81(3):420–6.

10. Mansour M, Leven E, Muellers K, Stone K, Mendu DR, Wajnberg A. Prevalence of SARS-CoV-2 Antibodies Among Healthcare Workers at a Tertiary Academic Hospital in New York City. J Gen Intern Med. 2020 Aug;35(8):2485–6.

11. Wantai : COVID-19 Serology and Molecular Tests [Internet]. Wantai BioPharm. [cited 2021 Sep 6]. Available from: http://www.ystwt.cn/covid-19/

12. Prevalence and Predictors of Stress, anxiety, and Depression among Healthcare Workers Managing COVID-19 Pandemic in India.pdf.

13. Iruretagoyena M, Vial MR, Spencer-Sandino M, Gaete P, Peters A, Delgado I, et al. Longitudinal assessment of SARS-CoV-2 IgG seroconversionamong front-line healthcare workers during the first wave of the Covid-19 pandemic at a tertiary-care hospital in Chile. BMC Infect Dis. 2021 May 26;21(1):478.

14. Murhekar MV, Bhatnagar T, Thangaraj JWV, Saravanakumar V, Kumar MS, Selvaraju S, et al. SARS-CoV-2 seroprevalence among the general population and healthcare workers in India, December 2020-January 2021. Int J Infect Dis IJID Off Publ Int Soc Infect Dis. 2021 May 19;108:145–55.

15. Edwards KM, Orenstein WA. COVID-19: Clinical features - UpToDate [Internet]. [cited 2021 Jul 20]. Available from: https://www.uptodate.com/contents/covid-19-clinical-features

16. Seroprevalence of antibodies against SARS-CoV-2 among health care workers in a large Spanish reference hospital | Nature Communications [Internet]. [cited 2021 Sep 6]. Available from: https://www.nature.com/articles/s41467-020-17318-x

17. CDC. Healthcare Workers [Internet]. Centers for Disease Control and Prevention. 2020 [cited 2021 Sep 6]. Available from: https://www.cdc.gov/coronavirus/2019-ncov/hcp/clinical-guidance-management-patients.html

18. Tabah A, Ramanan M, Laupland KB, Buetti N, Cortegiani A, Mellinghoff J, et al. Personal protective equipment and intensive care unit healthcare worker safety in the COVID-19 era (PPE-SAFE): An international survey. J Crit Care. 2020 Oct;59:70–5.

19. WHO-2019-nCoV-IPCPPE_use-2020.2-eng.pdf [Internet]. [cited 2021 Jul 20]. Available from: https://apps.who.int/iris/bitstream/handle/10665/331498/WHO-2019-nCoV-IPCPPE_use-2020.2-eng.pdf?sequence=1&isAllowed=y

20. Verbeek JH, Rajamaki B, Ijaz S, Sauni R, Toomey E, Blackwood B, et al. Personal protective equipment for preventing highly infectious diseases due to exposure to contaminated body fluids in healthcare staff. Cochrane Database Syst Rev. 2020 Apr 15;4:CD011621.

21. Hossain MA, Rashid MUB, Khan MAS, Sayeed S, Kader MA, Hawlader MDH. Healthcare Workers’ Knowledge, Attitude, and Practice Regarding Personal Protective Equipment for the Prevention of COVID-19. J Multidiscip Healthc. 2021 Feb 2;14:229– 38.

22. Agarwal, Ranjan P, Saraswat A, Kasi K, Bharadiya V, Vikram N, et al. Are health care workers following preventive practices in the COVID-19 pandemic properly? - A cross-sectional survey from India. Diabetes Metab Syndr [Internet]. 2021 [cited 2021 Jul 20];15(1):69–75. Available from: https://www.ncbi.nlm.nih.gov/pmc/articles/PMC7719197/

23. Gupta MK, Lipner SR. Personal protective equipment recommendations based on COVID-19 route of transmission. J Am Acad Dermatol. 2020 Jul 1;83(1):e45–6.

24. Protocol for assessment of potential risk factors for 2019-novel coronavirus (COVID-19) infection among health care workers in a health care setting [Internet]. [cited 2021 Jul 20]. Available from: https://www.who.int/publications-detail-redirect/protocol-for-assessment-of-potential-risk-factors-for-2019-novel-coronavirus-(2019-ncov)-infection-among-health-care-workers-in-a-health-care-setting

25. Palmore TN, Smith BA. COVID-19: Infection control for persons with SARS-CoV-2 infection - UpToDate [Internet]. [cited 2021 Jul 20]. Available from: https://www.uptodate.com/contents/covid-19-infection-control-for-persons-with-sars-cov-2-infection

26. Mick P, Murphy R. Aerosol-generating otolaryngology procedures and the need for enhanced PPE during the COVID-19 pandemic: a literature review. J Otolaryngol - Head Neck Surg J Oto-Rhino-Laryngol Chir Cervico-Faciale. 2020 May 11;49(1):29.

27. CoWIN Dashboard [Internet]. [cited 2021 Jul 20]. Available from: https://dashboard.cowin.gov.in/

28. Ritchie H, Ortiz-Ospina E, Beltekian D, Mathieu E, Hasell J, Macdonald B, et al. Coronavirus Pandemic (COVID-19). Our World Data [Internet]. 2020 Mar 5 [cited 2021 Jul 20]; Available from: https://ourworldindata.org/covid-vaccinations

29. Folegatti PM, Ewer KJ, Aley PK, Angus B, Becker S, Belij-Rammerstorfer S, et al. Safety and immunogenicity of the ChAdOx1 nCoV-19 vaccine against SARS-CoV-2: a preliminary report of a phase 1/2, single-blind, randomised controlled trial. The Lancet. 2020 Aug 15;396(10249):467–78.

30. Immune response to SARS-CoV-2 in health care workers following a COVID-19 outbreak: A prospective longitudinal study [Internet]. [cited 2021 Sep 6]. Available from: https://www.ncbi.nlm.nih.gov/pmc/articles/PMC7406471/

31. Sharma P, Chawla R, Bakshi R, Saxena S, Basu S, Bharti PK, et al. Seroprevalence of antibodies to SARS-CoV-2 and predictors of seropositivity among employees of a teaching hospital in New Delhi, India. Osong Public Health Res Perspect. 2021 Apr;12(2):88–95.

32. Kantele A, Lääveri T, Kareinen L, Pakkanen SH, Blomgren K, Mero S, et al. SARS-CoV-2 infections among healthcare workers at Helsinki University Hospital, Finland, spring 2020: Serosurvey, symptoms and risk factors. Travel Med Infect Dis [Internet]. 2021 Jan 1 [cited 2021 Jul 20];39. Available from: https://covid19.elsevierpure.com/en/publications/sars-cov-2-infections-among-healthcare-workers-at-helsinki-univer

33. Klevebro S, Bahram F, Elfström KM, Hellberg U, Hober S, Merid SK, et al. Risk of SARS-CoV-2 exposure among hospital healthcare workers in relation to patient contact and type of care. Scand J Public Health. 2021 Jun 19;14034948211022434.

34. Iruretagoyena M, Vial MR, Spencer-Sandino M, Gaete P, Peters A, Delgado I, et al. Longitudinal assessment of SARS-CoV-2 IgG seroconversionamong front-line healthcare workers during the first wave of the Covid-19 pandemic at a tertiary-care hospital in Chile. BMC Infect Dis. 2021 Dec;21(1):478.

35. Müller K, Girl P, Ruhnke M, Spranger M, Kaier K, von Buttlar H, et al. SARS-CoV-2 Seroprevalence among Health Care Workers—A Voluntary Screening Study in a Regional Medical Center in Southern Germany. Int J Environ Res Public Health. 2021 Apr 8;18(8):3910.

36. Lumley SF, Wei J, O’Donnell D, Stoesser NE, Matthews PC, Howarth A, et al. The Duration, Dynamics, and Determinants of Severe Acute Respiratory Syndrome Coronavirus 2 (SARS-CoV-2) Antibody Responses in Individual Healthcare Workers. Clin Infect Dis. 2021 Aug 1;73(3):e699–709.

37. Milazzo L, Lai A, Pezzati L, Oreni L, Bergna A, Conti F, et al. Dynamics of the seroprevalence of SARS-CoV-2 antibodies among healthcare workers at a COVID-19 referral hospital in Milan, Italy. Occup Environ Med. 2021 Aug 1;78(8):541–7.

38. Hains DS, Schwaderer AL, Carroll AE, Starr MC, Wilson AC, Amanat F, et al. Asymptomatic Seroconversion of Immunoglobulins to SARS-CoV-2 in a Pediatric Dialysis Unit. JAMA. 2020 Jun 16;323(23):2424–5.

39. Chen MIC, Lee VJM, Barr I, Lin C, Goh R, Lee C, et al. Risk Factors for Pandemic (H1N1) 2009 Virus Seroconversion among Hospital Staff, Singapore. Emerg Infect Dis. 2010 Oct;16(10):1554–61.

40. Hunter BR, Dbeibo L, Weaver CS, Beeler C, Saysana M, Zimmerman MK, et al. Seroprevalence of severe acute respiratory coronavirus virus 2 (SARS-CoV-2) antibodies among healthcare workers with differing levels of coronavirus disease 2019 (COVID-19) patient exposure. Infect Control Hosp Epidemiol. 2020 Dec;41(12):1441–2.

